# The origin of the MV Hondius Andes outbreak

**DOI:** 10.64898/2026.11.04.26366130

**Authors:** Lissette Ulloa-Zepeda, Edyth Parker, Karthik Gangavarapu, Andrés Castillo, Catalina Pavez, Jorge Fernandez, Marcelo Vilches, Pablo Vial, Andrew Rambaut, Kristian G. Andersen, Cecilia Vial

## Abstract

In April-July 2026, Andes virus (ANDV) caused an outbreak aboard the MV Hondius cruise ship resulting in 12 confirmed cases and three deaths. Before symptom onset, the index case had travelled through hantavirus-endemic regions of Chile and Argentina, leaving the geographic origin of infection unresolved. We conducted phylogenetic and phylogeographic analyses using newly generated ANDV genomes from human cases and rodent reservoirs to investigate the likely origin of the outbreak. Through phylogenetic analyses, we show that the outbreak genome falls within a geographically structured lineage spanning southern Chile and Argentine Andean Patagonia. Using phylogeographic reconstructions across all three segments, integrated with the index case’s travel itinerary, we identified the Toltén-Lanco area of southern Chile as the most likely geographic origin of the outbreak genome, with two candidate locations accounting for >98% of the posterior support. However, the index case visited this area 62-66 days before symptom onset, exceeding the conventionally reported ANDV incubation period of 7-42 days. Thus, while genomic evidence supports a southern Chilean origin of the outbreak genome, the timing and circumstances of transmission to the index case remain unresolved.

## Main Text

On 2 May 2026, a cluster of severe acute respiratory illness aboard the MV Hondius cruise ship involving two deaths and a third critically ill passenger was reported to the World Health Organization (WHO)^1,2^. The index case had developed nonspecific symptoms on 3 April and succumbed to the virus aboard the vessel on 11 April^3^. Hantavirus infection was confirmed on 2 May and identified as Andes virus (ANDV; *Orthohantavirus andesense*) on 5 May^3–5^. Subsequent human-to-human transmission aboard the ship resulted in thirteen cases within eight weeks, with a case fatality rate of 23%^6^. This outbreak highlights the epidemic potential of ANDV, the only Hantavirus known to transmit from human-to-human following zoonotic spillover^7–9^.

The index and secondary cases were a couple who had spent five months traveling across South America before boarding the cruise ship on 1 April^1^ in Ushuaia, Tierra del Fuego, Argentina. Their travel route was well documented and reconstructed from their observation records, geolocated through the eBird birdwatching database^10^, and from national government agencies^11^. The travel itinerary included extensive travel through Argentina and Chile, including Argentine Patagonia (mid-December 2025 to early January 2026), southern Chile (January) and central Chile (Maule region, Chile; mid-February), northeastern Argentina (late February and mid-March), and Uruguay (late March) (Fig.1a). They returned to Argentina on March 27 before travelling to Ushuaia where the MV Hondius departed. This itinerary traversed a large swathe of the ANDV-endemic range in Chile and Argentina where exposure to ANDV is plausible given the broad distribution of the principal reservoir host, the long-tailed pygmy rice rat (*Oligoryzomys longicaudatus*), from the southern Atacama Desert in Chile (∼27°S) to the Tierra del Fuego archipelago (∼54°S)^12,13^. Their route also encompassed regions associated with other members of *Orthohantavirus andesense* species, including the Lechiguanas and Orán viruses^14,15^.

The index case’s extensive travel history, together with the broad host distribution and the geographic range of known ANDV circulation provide multiple plausible sites of zoonotic exposure. Here, we investigated the possible hypotheses for the geographic origin of the MV Hondius outbreak (Table 1): exposure in (H1) the southern Chile region (“Southern”), (H2) central Chile (“Central”), (H3) Argentine Norte Grande (“Norte Grande”); or (H4) Ushuaia (“Tierra del Fuego”) immediately before boarding the cruise ship. We investigated these hypotheses by integrating epidemiological evidence with publicly available *O. andesense* genomic data and newly generated ANDV sequences from south and central Chile.

**Table 1.** Competing exposure hypothesis for the MV Hondius outbreak origins.

| Hypothesis | Candidate exposure region | Travel period | Time before symptoms onset (days) | Epidemiological context |
|---|---|---|---|---|
| H1 | Southern Chile | 8 - 31 Jan | 62 - 85 | ANDV-endemic region; documented circulation and reservoir host present |
| H2 | Central Chile | 12 - 16 Feb | 46 - 50 | ANDV-endemic region; documented circulation and reservoir host present |
| H3 | Argentine Norte Grande | 20 Feb - 13 Mar | 21 - 42 | Other *O. andesense* virus-endemic areas (e.g. Lechiguanas and Orán viruses); presence of reservoir host not documented |
| H4 | Ushuaia, Tierra del Fuego | 27 Mar - 1 Apr | 2 - 7 | ANDV reservoir host present; no reported human hantavirus cases |

We first characterized the phylogenetic relationship of the outbreak genomes to the broader *O. andesense* diversity. Prior to the MV Hondius outbreak, we had established a program to strengthen genomic surveillance of ANDV in Chile^16^. As part of this effort, in collaboration with the Chilean Public Health Institute, we assembled a genomic dataset comprising previously and newly generated sequences from ten rodent and nineteen human samples collected from 2011 to 2026 across central (Metropolitan, O’Higgins and Maule regions), southern (Ñuble, Bíobio, La Araucanía, Los Ríos and Los Lagos regions) and austral (Aysén) regions (Extended Data Fig.1). The dataset included 12 L, 15 M, and 28 S-segment sequences, which correspond to the RNA-dependent RNA polymerase (L segment), glycoproteins (M segment) and nucleoprotein (S segment) genes (Supplementary Table 1).

**Fig. 1.**
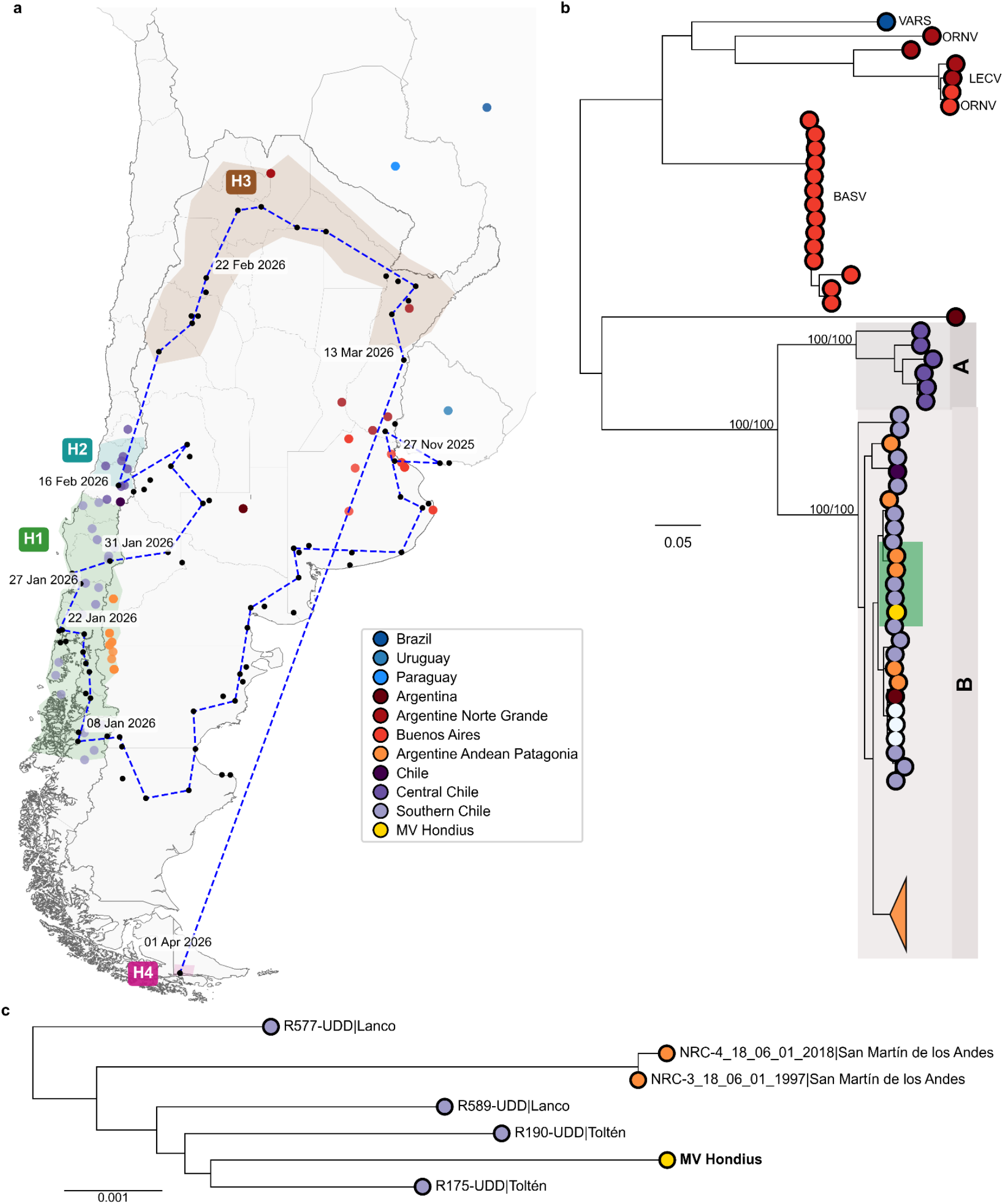
Epidemiological context and phylogenetic relationship of the MV Hondius outbreak genome. **a,** Geographic distribution of *O. andesense* sequences included in the analyses and travel itinerary of the index case. Colored circles indicate sequence sampling locations, black circles connected by blue dashed lines indicate the documented travel itinerary of the index case; approximated dates are shown along the route. Shaded areas and labels (H1-H4) highlight the geographic and temporal portions of the itinerary corresponding to the exposure hypotheses considered in this study. **b,** Maximum-likelihood phylogeny reconstructed for the M segment. Tips are colored according to geographic sampling location, and the MV Hondius outbreak genome is highlighted in yellow. Within the ANDV lineage, two major geographically structured lineages central (A) and southern (B) are shaded in gray, while B1.2.2 lineage containing the outbreak genome is highlighted in green. Other hantaviruses included are labeled as LECV (Lechiguanas virus), ORNV (Orán virus) and BASV (Buenos Aires virus). Bootstrap support values are shown for selected nodes. **c,** Detail view of B.1.2.2 lineage containing the outbreak genome and its closest sampled relatives.

We used a representative sequence for each genomic segment, with variable sites masked, to assess the evolutionary relationship between the outbreak genome and the broader *O. andesense* diversity. We reconstructed maximum-likelihood (ML) phylogenies independently for each segment using publicly available genomes. Across all three segments, the MV Hondius outbreak genome consistently clustered within the ANDV lineage spanning Chile and adjacent Argentinean provinces as expected (Fig.1b, Extended Data Fig.2)^17^. Viruses sampled outside this geographic range fall within distinct *O. andesense* lineages, arguing against H3 (Norte Grande) under the currently sampled viral diversity, whereas H4 (Tierra del Fuego) could not be directly evaluated due to lack of available sequences from this region.

**Fig. 2.**
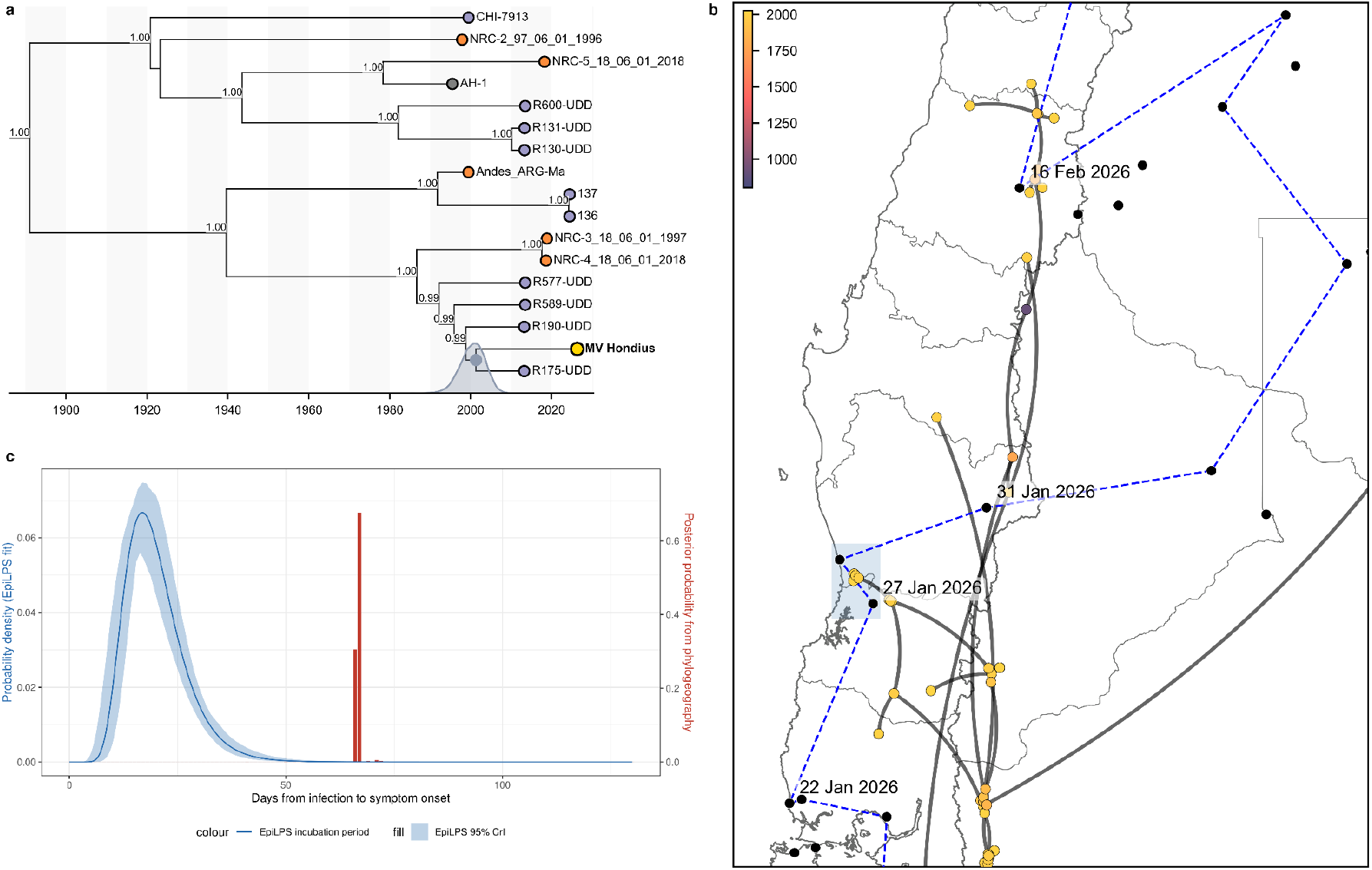
Bayesian phylogeographic reconstruction of the MV Hondius outbreak lineage for the M segment. **a**, Time-resolved maximum clade credibility subtree containing the MV Hondius outbreak genome. The outbreak lineage is highlighted in yellow (PP > 0.99 are shown). **b,** Continuous phylogeographic reconstruction for the L segment within the geographic region relevant to the index case travel history. Grey lines represent inferred lineage dispersal trajectories, and colored points indicate timing of viral dissemination. Black circles connected by blue dashed lines show the travel itinerary with dates indicated along the route. **c,** Overlap between incubation-period distribution estimated from documented rodent-to-human transmission and the incubation period inferred from the phylogeographic reconstruction. The blue curve shows the incubation period distribution fitted with EpiLPS using rodent-to-human transmission data (Vial et. al)^24^ with the shaded area indicating the 95% credible interval. Red bars show the probability of incubation periods inferred from the phylogeographic analysis.

Following the Pango lineage nomenclature framework^18^, we designated two major geographically structured subclades within the ANDV lineage spanning Chile and adjacent Argentinean provinces: lineage A, comprising sequences from central Chile, spanning the Metropolitan to the Maule region (∼33 and 36°S), and lineage B, comprising sequences from southern Maule to the Argentine Andean Patagonia (46°S and 71°W) (Fig 1b, Extended Data Fig.2). We found that the outbreak genome clustered within lineage B with strong support across all segments (Fig.1c and Extended Data Fig.3a-c). Specifically, the outbreak genome is nested within a well-supported clade (bootstrap support > 98) that consistently includes rodent-derived sequences from Toltén (La Araucanía region, Chile) and Lanco (Los Ríos region, Chile), located approximately 48 km apart, and human-derived sequences from San Martín de los Andes (Neuquén, Argentina), previously reported as cases epidemiologically unrelated to the 2018-19 Epuyén outbreak (NRCs)^8^, which is ∼150km east across the Andes and the Chile-Argentina border (Extended Data Fig.4). This well-supported trans-Andean clade spans southern Chile and northwestern Argentine Andean Patagonia and is hereafter referred to as lineage B.1.2.2 (Extended Data Fig.3a-c). This phylogenetic placement supports H1 (Southern) that the index case was infected while traveling from the Aysén to La Araucanía regions between 8 and 31 January. Further, given that the outbreak genome does not cluster within the central Chilean lineage A, it provides no support for H2 (Central).

Within lineage B.1.2.2 the outbreak genome was most closely related to our sequences R175 and R190, both sampled from rodents captured in Toltén (39°17’S) in 2013 (Fig.1c and Extended Data Fig.4). Despite minor variations in the tree topology within the lineage B.1.2.2, with R175 and R190 consistently representing the closest relatives, the phylogenetic congruence across segments indicates no evidence of reassortment involving the outbreak genome (Extended Data Fig.3d). The outbreak genome and their closest rodent-derived relatives (Toltén R175/R190) differed by 47, 23 and 6 SNPs across the L, M, and S segment coding sequences, respectively. Meanwhile, viruses sampled in Lanco differed from the outbreak genome by 59, 29 and 12 SNPs on average, whereas those from San Martín de los Andes in Argentina differed by 80, 35 and 17 SNPs, respectively.

Given their genetic divergence from the outbreak genome, we performed a Bayesian phylogenetic reconstruction to investigate whether the Toltén viruses shared a recent common ancestor with the outbreak genome. We found that the outbreak genome and the Toltén sequences shared a common ancestor over two decades before the 2026 outbreak, with overlapping time to the most recent common ancestor (tMRCA) estimates across segments (Fig.2a and Extended Data Fig.5a-c). The median tMRCA was estimated at November 2000 95% HPD: [October 1995, October 2005]) for the L segment, August 2000 (95% HPD: [August 1994, April 2006]) for the M segment, and November 2003 (95% HPD: [January 1996, July 2010]) for the S segment (Extended Data Fig.5a-c). Thus, although the Toltén viruses represent the closest sampled relatives, they do not share a recent ancestor. The prolonged period of divergence between the outbreak sequence and its closest relatives likely reflects the sparse genomic sampling of ANDV in the region, restricting the ability to constrain uncertainty in divergence times^19,20^.

Our ML phylogenetic analyses support H1 (Southern), associating the MV Hondius outbreak genome with viruses circulating in southern Chile-Argentine Andean Patagonia, where the index case traveled between 8 and 31 January. However, this visit occurred 62-85 days before symptoms onset, well outside the documented ANDV incubation period of 7-42 days (Table 1). To further resolve the geographic and temporal origin of the outbreak and investigate this conundrum—while accounting for the associated phylogenetic uncertainty—we performed Bayesian continuous phylogeographic reconstructions of the ANDV lineage. We sampled candidate geographic origins of the outbreak genome from locations visited by the index case during their reconstructed travel (Fig.1a). Consistent with our ML phylogenetic analyses (Fig. 1b) and H1 (Southern), we found that the outbreak genome was placed within lineage B.1.2.2 across all three segments, clustering with both Toltén sequences (R175/R190) with high posterior support (posterior probability >0.99) (Extended data fig. 5a-c). Across all three segments, two candidate locations for the geographic origin of the outbreak genome consistently accounted for >98% of the posterior probability: one approximately 20km south of Toltén and the other approximately 20km west of Lanco (Extended Data Fig.5d-f). Together these locations represent an area of approximately 2,800 km^2^ encompassing the Toltén-Lanco area (Fig. 2b).

The genomic evidence, based on both our ML (Fig. 1b) and Bayesian (Fig. 2a, b) phylogenetic analyses, supports a southern Chile origin consistent with H1 (Southern). Further, our phylogeographic inference specifically identifies the Toltén-Lanco area in southern Chile as the most likely origin of the outbreak. According to the reconstructed travel itinerary, the index case visited this area in late January, 62-66 days before symptoms onset, within the broader 62–85-day interval corresponding to travel across southern Chile under H1. This more geographically specific scenario would still imply an incubation period substantially longer than the documented 7-42 days for ANDV (Table 1). To determine where a putative incubation-period of 62-66 days falls within the expected distribution for ANDV, we used the EpiLPS package^21^ to estimate the incubation period for rodent-to-human transmission using data from a study of 20 patients with documented rodent exposure and symptom onset dates^15^ (Fig. 2c). EpiLPS compares classic parametric distributions commonly used to model incubation periods lognormal, gamma, and Weibull, alongside a flexible spline model^22^. We selected the lognormal distribution based on the lowest Bayesian information criterion, with an estimated mean incubation period of 20 days (95% credible interval [CrI]: 18.1-21.4). To assess whether the lognormal distribution adequately described the data, we also fitted it directly to the interval-censored incubation data using maximum likelihood estimation with the R package fitdistrplus^23^. We found that an incubation period consistent with exposure during the Toltén-Lanco visit was highly unlikely based on the estimated incubation-period distribution from previously documented zoonotic transmission events, with negligible overlap between the incubation period distribution estimated from documented rodent-to-human transmission events and that inferred from the phylogeographic analysis (overlap coefficient = 6.6e-04 (95% CrI = [3.7e-04, 1.4e-03]), Fig. 2c).

Together, these findings reveal conflicting evidence across the proposed exposure hypotheses (Table 2). As outlined, H1 (Southern) is supported by the genomic evidence, but is inconsistent with known incubation periods^24^. Potential exposure in central Chile (H2) would have occurred closer to, but still outside of, the standard incubation window. However, this hypothesis is not supported by the phylogenetic analyses as the evidence shows that lineage B is unlikely to circulate in this region. The documented incubation period for ANDV of 7-42 days suggests a likely exposure window from February 20 to March 27. At this time, the index case traveled through the Argentine Norte Grande region (H3) including the Salta and Corrientes provinces, more than 1,700km from the Toltén-Lanco area. However, available sequences sampled from these regions belong to other *O. andesense* viruses rather than to the ANDV lineage containing the outbreak genome. Finally, potential exposure in Ushuaia (H4) would have occurred only 2-7 days before symptom onset and would require a short incubation period. Further, although recent reports indicate hantavirus detection in rodent reservoirs in Tierra del Fuego, with preliminary evidence suggesting genetic differences from the outbreak genome, no human cases have been reported in the region^25,26^. Therefore, although the geographic range of the reservoir makes local circulation possible, there is currently no epidemiological or genomic evidence supporting H4 (Tierra del Fuego).

**Table 2.**
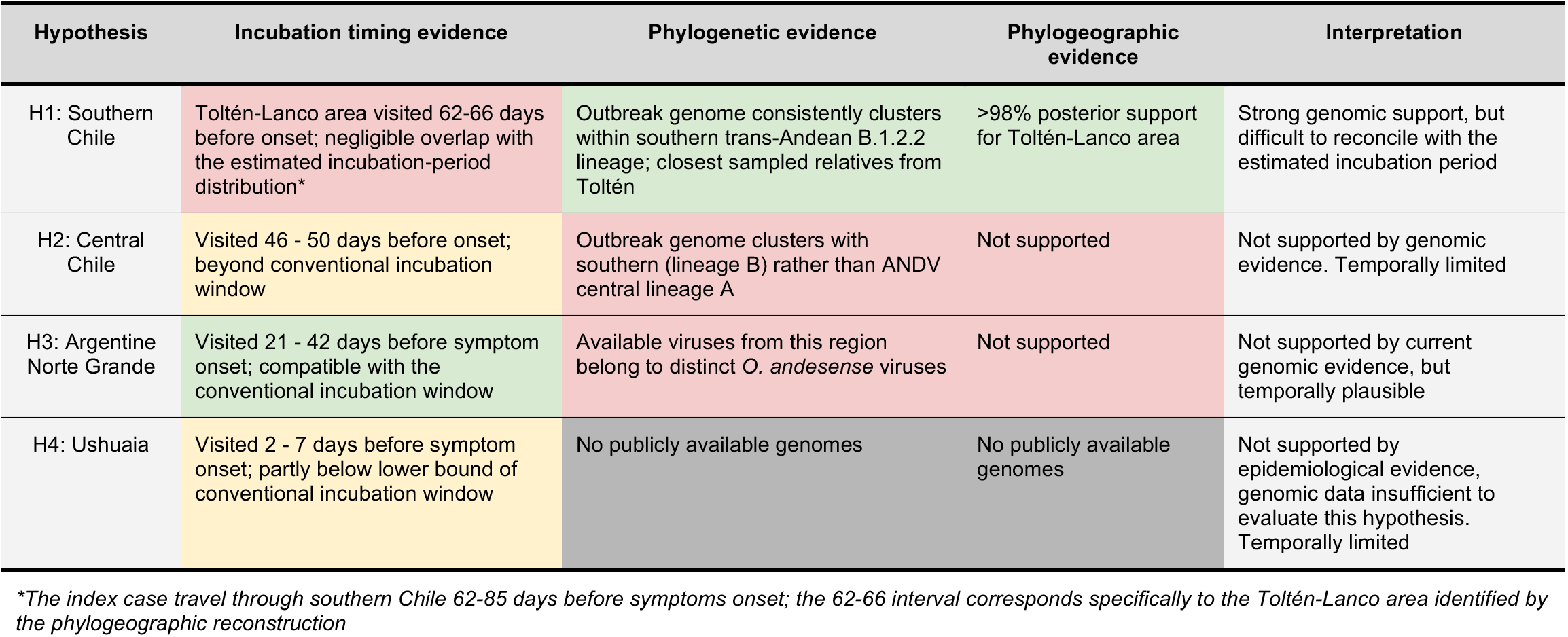
Evaluation of the proposed exposure hypotheses based on genomic and epidemiological evidence.

| **Hypothesis** | **Incubation timing evidence** | **Phylogenetic evidence** | **Phylogeographic evidence** | **Interpretation** |
|---|---|---|---|---|
| H1: Southern Chile | Toltén-Lanco area visited 62-66 days before onset; negligible overlap with the estimated incubation-period distribution* | Outbreak genome consistently clusters within southern trans-Andean B.1.2.2 lineage; closest sampled relatives from Toltén | >98% posterior support for Toltén-Lanco area | Strong genomic support, but difficult to reconcile with the estimated incubation period |
| H2: Central Chile | Visited 46 - 50 days before onset; beyond conventional incubation window | Outbreak genome clusters with southern (lineage B) rather than ANDV central lineage A | Not supported | Not supported by genomic evidence. Temporally limited |
| H3: Argentine Norte Grande | Visited 21 - 42 days before symptom onset; compatible with the conventional incubation window | Available viruses from this region belong to distinct *O. andesense* viruses | Not supported | Not supported by current genomic evidence, but temporally plausible |
| H4: Ushuaia | Visited 2 - 7 days before symptom onset; partly below lower bound of conventional incubation window | No publicly available genomes | No publicly available genomes | Not supported by epidemiological evidence, genomic data insufficient to evaluate this hypothesis. Temporally limited |

*\*The index case travel through southern Chile 62-85 days before symptoms onset; the 62-66 interval corresponds specifically to the Toltén-Lanco area identified by the phylogeographic reconstruction*

Overall, none of the exposure scenarios for the origin of the MV Hondius outbreak reconciles the genomic and epidemiological evidence. A major limitation in resolving these scenarios is the sparse genomic sampling of ANDV. Although the Toltén viruses are the closest relatives of the outbreak genome, their common ancestor predates the outbreak by more than two decades, indicating substantial unsampled viral diversity in this region. The geographic distribution of B.1.2.2 may therefore be incompletely characterized, encompassing locations visited more recently by the index case, within the documented incubation period. Although incomplete sampling prevents this possibility from being excluded, the strong geographic structure across the *O. andesense* phylogenies suggests that such a scenario would require circulation across substantial geographic distances and major ecological and topographic transitions (Fig. 1a, b)^12,13^. Such landscape features can constrain viral dispersal and shape the geographic distribution of lineages, as illustrated by previous studies of ANDV^27^, LASV^28,29^ and MPXV^30^ highlighting the influence of environmental factors on viral circulation.

Alternatively, documented ranges for ANDV’s incubation period may have incompletely captured its full distribution. A few studies with relatively well-defined exposure windows have reported incubation periods of up to 39-40 days, whereas a 42-day period has been accepted for ANDV contact surveillance^9,24^. Some public health guidance reports even longer periods, but only up to 45 days^31^. Incubation-period estimates for sporadic zoonotic transmission events are inherently difficult to report as the precise timing of exposure is rarely known and individuals may experience multiple potential exposure events^24^. Reported estimates may therefore incompletely capture the full distribution, particularly its upper (right) tail^32,33^. Although our analysis indicates that an incubation period consistent with exposure during the Toltén-Lanco visit in January is highly unlikely, uncertainty in the upper tail means that an unusually long incubation period cannot be excluded. Such an extended incubation period could reconcile the timing of H1 (Southern) with the strong genomic evidence supporting this scenario. Whether differences in infectious dose could contribute to unusually prolonged incubation periods in ANDV infections remains unknown.

Lastly, discordance between the genomic and epidemiological evidence may indicate that exposure did not occur through direct contact with a locally infected rodent at one of the locations considered in our four hypotheses (Table 1). Other scenarios could include exposure to an infected rodent or contaminated material transported between locations, potentially involving the vehicle used while traveling through endemic regions^34^, or transmission from an unidentified human case infected earlier in the travel itinerary. Previous work on the Sin Nombre virus has demonstrated the value of genomic evidence for distinguishing among geographically separated exposure sites when conventional epidemiological evidence is inconclusive^35^. However, unlike those cases, where matching human and rodent-derived viral sequences helped identify probable exposure sites, the MV Hondius outbreak presents a substantial discrepancy between the genomic evidence and timing of potential exposure. These alternative scenarios cannot be resolved with the available genomic and epidemiological data but could potentially decouple the geographic origin of the outbreak genome from the location where transmission to the index case occurred.

Despite these uncertainties, our findings provide strong genomic support for a southern Chilean origin of the MV Hondius outbreak, while the precise origin of the outbreak remains unresolved and should be interpreted cautiously pending additional genomic and epidemiological data. Denser genomic surveillance of the animal reservoir across southern Chile and Argentina will be essential to characterize unsampled viral diversity, while improved characterization of the ANDV incubation-period distribution will help resolve the temporal uncertainty surrounding potential exposure events.

---

**Extended Data Figure 1.**
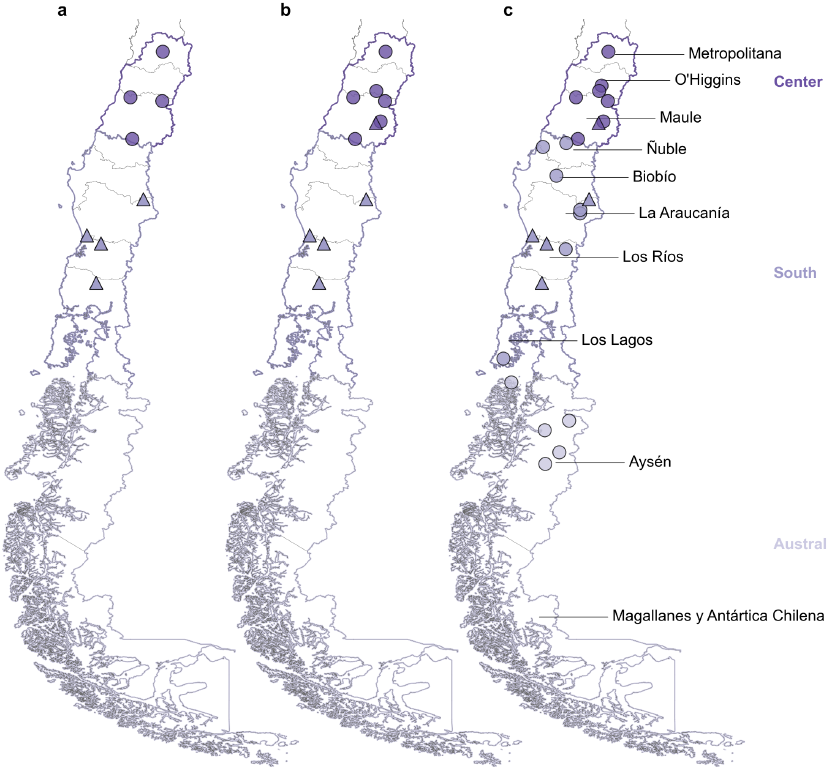
Sampling locations of the new ANDV sequences used in this study. Symbols represent host type (triangle = rodents and circles = human); color indicates the corresponding sampling macrozones for (a) L, (b) M and (c) S segments.

**Extended Data Figure 2.**
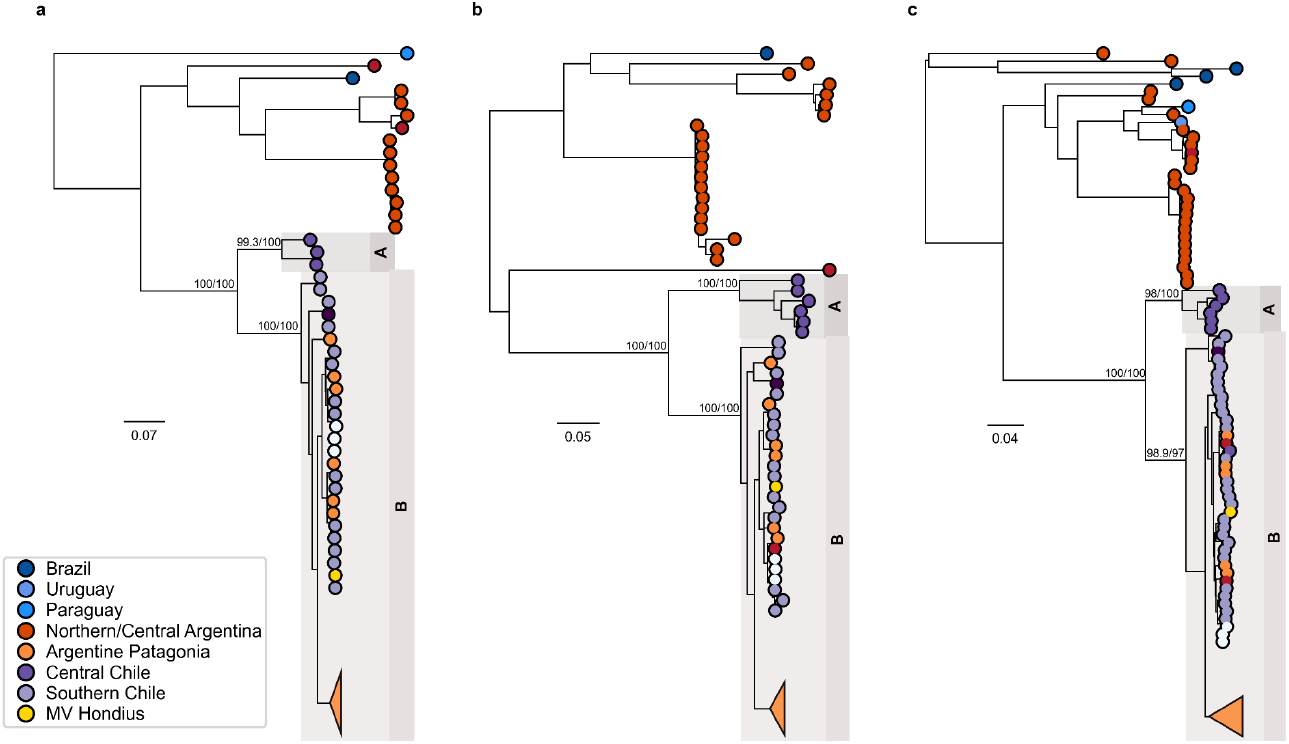
Maximum-likelihood phylogenies of the L, M and S segments of *O. andesense*. Maximum-likelihood phylogenies were independently reconstructed for the L (**a**), M (**b**) and S (**c**) segments of sequences included in this study. The two major geographically structured ANDV lineages, central (A) and southern (B) are highlighted in each tree. Node labels indicate SH-aLRT/UFBoot support values for selected nodes. Tips colors indicate the geographic sampling location of each sequence, with outbreak genome highlighted in yellow. Scale bars indicate nucleotide substitutions per site.

**Extended Data Figure 3.**
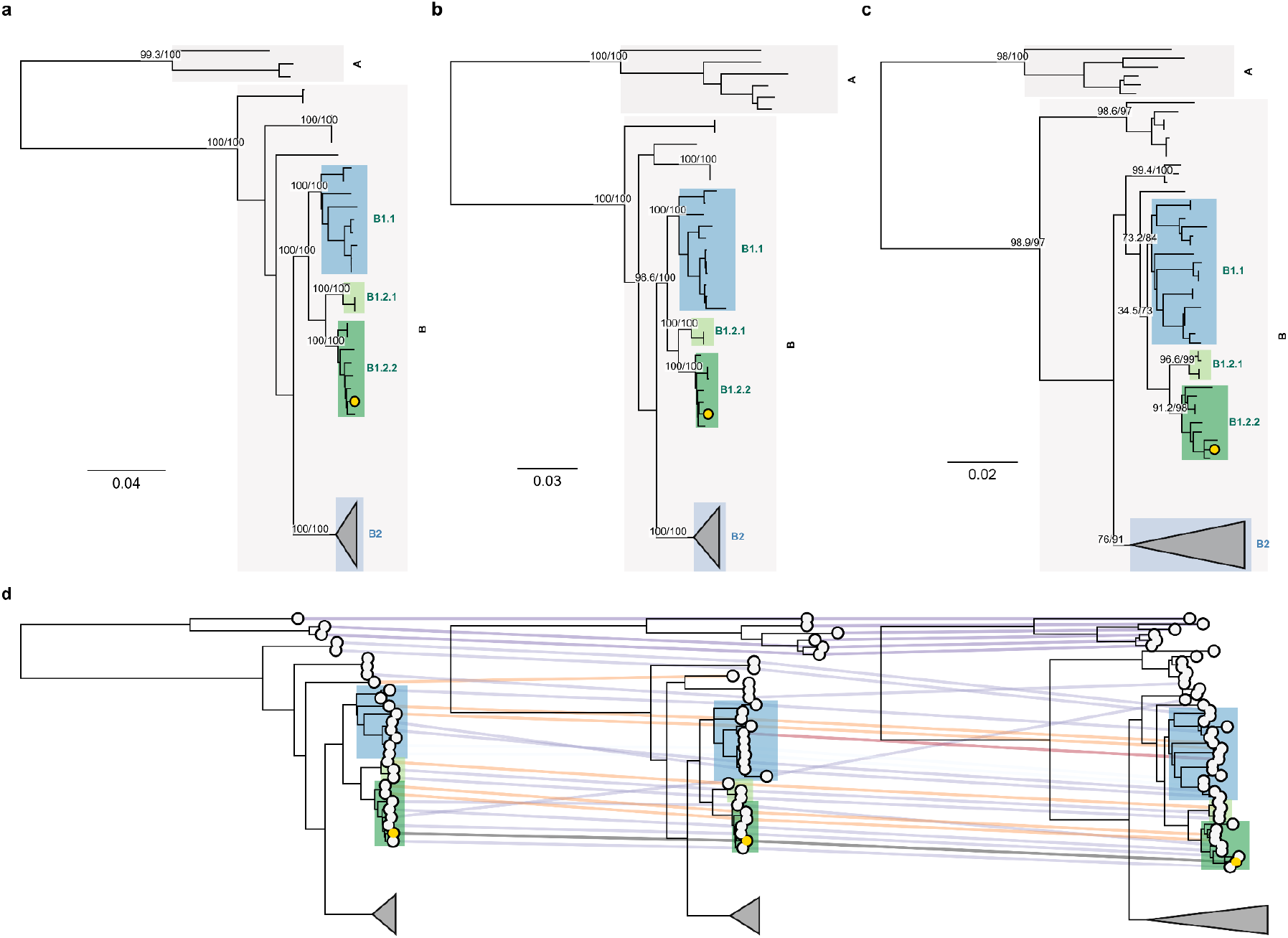
Maximum likelihood reconstruction of ANDV lineage complete genome phylogenies. Phylogenetic trees for the (**a**) L, (**b**) M and (**c**) S segment of ANDV. Sublineages are highlighted in each tree and its statistical support (SH-aLRT and UFB) is annotated. Scale bars indicate the number of nucleotide substitutions per site. **d**, Tanglegram of maximum-likelihood phylogenies of L, M and S ANDV segments. Lines connect tips of a single sampled genome across each segment, colored by sampling location.

**Extended Data Figure 4.**
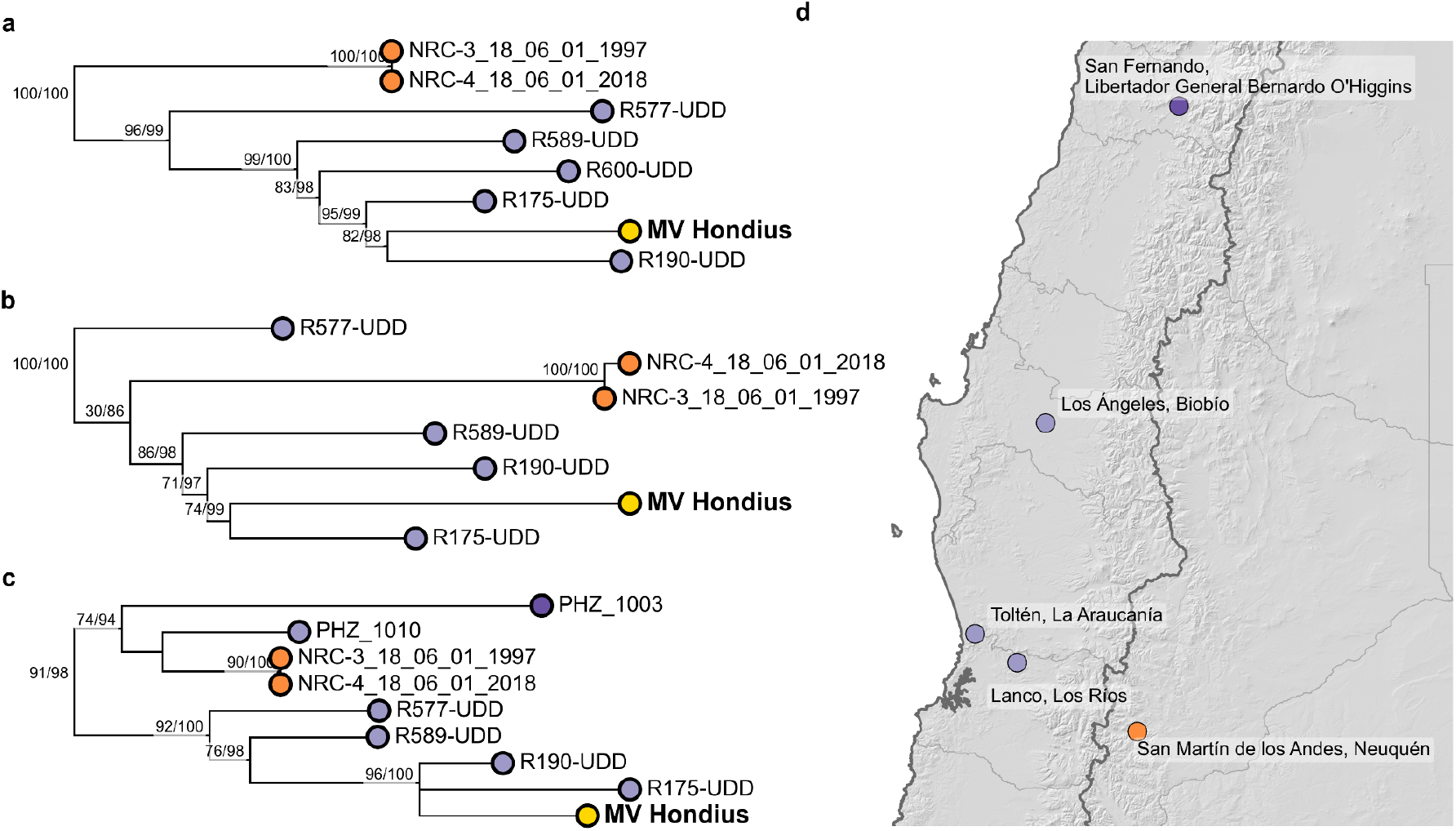
Detailed phylogenetic relationships of the outbreak genome across segments. **a-c**, ML phylogenies for the L (**a**), M (**b**) and S (**c**) segments. The outbreak sequence is highlighted in yellow, and the remaining tips are colored according to sampling location. Node labels indicate SH-aLRT/UFBoot support values. **d**, Map showing the sampling locations of the sequences included in the highlighted clade.

**Extended Data Figure 5.**
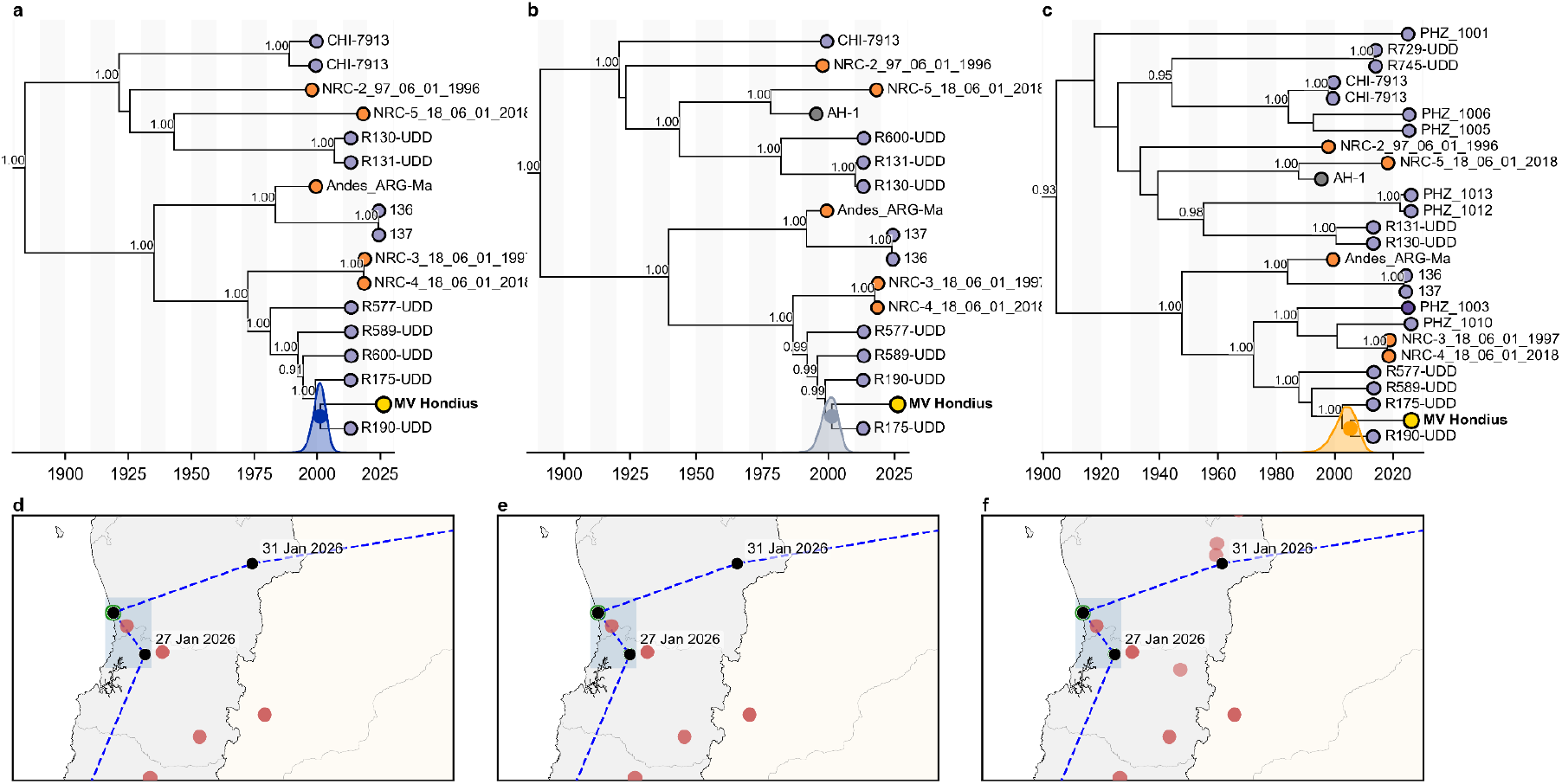
Bayesian phylogenetic and inferred geographic origin of the outbreak genome across segments. Maximum clade credibility (MCC) trees inferred for the (**a**) L, (**b**), M, and (**c**), S segments, of the outbreak genome and its closest sampled relatives. Shaded distribution indicates the estimated time to the most recent common ancestor (tMRCA) of the outbreak genome and Toltén sequences, with the corresponding 95% highest posterior density (HPD) interval. Node labels indicate posterior probabilities. **b-f**, inferred geographic origin of the outbreak lineage shown in relation to the documented travel itinerary, with approximate dates shown along the route. Green points indicate the two possible geographic origin locations of the outbreak lineage that accounted for >98% of the posterior probability, and the shaded area represents the uncertainty.

**Extended Data Figure 6.**
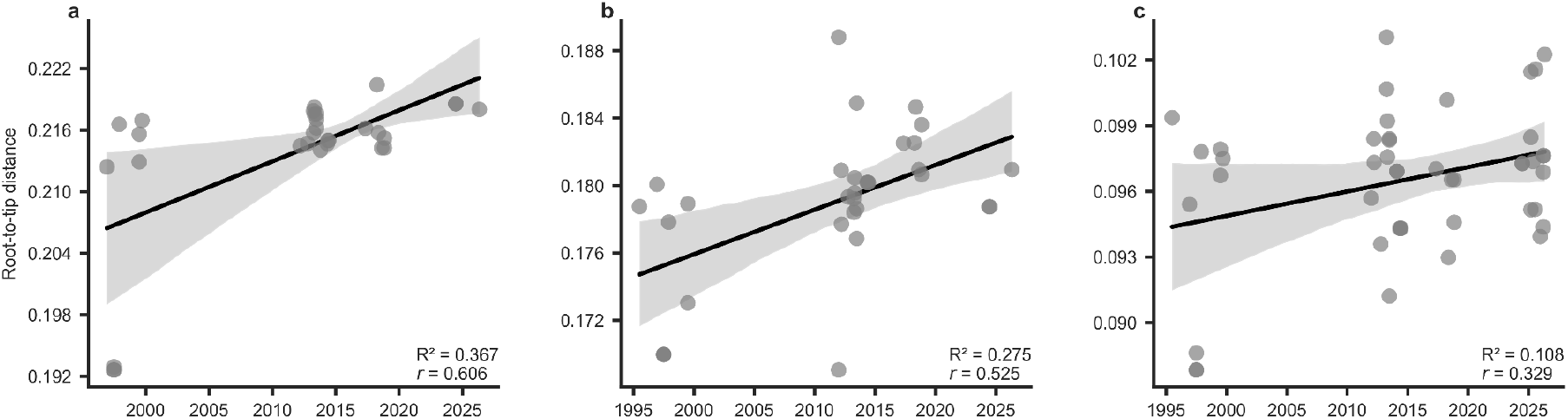
Temporal regression of the ANDV phylogeny. Each point represents a sequence’s divergence from the root in time for the (**a**) L, (**b**) M and (**c**) S segment dataset.

## ONLINE METHODS SECTION

### Ethics declaration

This study was developed collaboratively by a partnership among the Hantavirus and Zoonosis program at Universidad del Desarrollo Clínica Alemana (Chile), the Chilean Public Health Institute, the Scripps Research Institute (USA) and the Institute of Evolutionary Biology at The University of Edinburgh (Scotland).

All human subject participation was in compliance with the Declaration of Helsinki and Chilean Regulation (Vial et al, 2015). All participants signed an informed consent, previously approved by institutional ethics committees.

### Rodent sample collection

Rodent samples included in this study were obtained through field sampling conducted in south-central Chile where confirmed HCPS cases had been reported. Rodent trapping, sample collection and biosafety procedures have been described in detail elsewhere^16^. Briefly, rodents were captured across Metropolitana and Los Lagos regions during 2011-2014, and organs (heart, kidney, spleen, liver and/or lung) from seropositive specimens were collected and stored. Viral RNA was detected from positive serology specimens and sequenced through Oxford Nanopore Technology platform.

### Patient sample collection

The dataset was generated through collaborative efforts among the Chilean Public Health Institute and the Hantavirus and Zoonosis Program between 2011 and 2020. Whole-blood samples were collected by venipuncture from RT-qPCR confirmed HCPS cases between days 1 and 5 of hospitalization. Samples were obtained either from patients enrolled in research projects approved by the Ethics Committee of the Faculty of Medicine at Clínica Alemana Universidad del Desarrollo, or through the national hantavirus surveillance program as part of the epidemiological surveillance carried out by the Ministry of Health through the Reference Laboratory of the Public Health Institute of Chile.

### Nucleic acid extraction

Viral RNA was extracted from plasma and buffy-coat using the QIAamp Viral RNA Mini Kit (Qiagen), according to the manufacturer’s instructions. Viral RNA was tested by RT-qPCR targeting the S segment of the ANDV genome as described in Vial et al. 2016^36^.

### One-Step RT-PCR amplification and genomic sequencing

Reverse transcription and amplification of viral RNA were performed using a SuperScriptTM IV one-step RT-PCR system (Invitrogen), and the library preparation was performed using Illumina CovidSeq Test (Illumina, San Diego, CA, USA), both protocols are described in Ulloa-Zepeda et al. 2026^16^.

### Genome assembly

Raw sequencing reads were processed with FastP (v0.23.2)^37^ for trimming adapters and filtering low-quality bases. Genome reconstruction was performed using different workflows according to the sequencing platform. Illumina reads were either assembled de novo using metaSPAdes(v3.15.5)^38^ when no suitable reference genome was available or mapped to a reference (L: NC_003468, M: NC_003467 and S: NC003466) as described elsewhere (Ulloa-zepeda, 2026). Briefly, reads were mapped using BWA MEM (0.7.17.r1198)^39^, primers were trimmed using iVar trim (v1.4.2)^40^ followed by consensus calling with samtools (v1.16.1)^41^ and iVar consensus. High-quality long-reads from ONT platform were filtered with Porechop (v0.2.4)^42^ and NanoFilt (v2.8.0)^43^, mapped to a reference genome with Minimap2 (v2.24-r1122)^44^, primer sequences were trimmed using iVar and consensus was generated samtools and iVar.

A single MV Hondius outbreak genome (PP_006WBLH.2)^4^ was used to represent the outbreak but to remove variation that had arisen within the outbreak, any sites known to be variable amongst the other outbreak sequences were masked as N^45^.

### Phylogenetic analyses

The phylogenetic placement of the outbreak-associated genome was first evaluated in a broader species context. For each segment, all publicly available O. andesense sequences from Pathoplexus^46^ covering > 75% were included. All synthetic sequences were removed from the dataset (derived from constructs, passaged or patent sequences). We aligned the sequences using Mafftv7.505^47^ under the automatic selection and trimmed the 5′ and 3′ ends. We reconstructed the complete O. andesense phylogeny using IQ-Tree2 v2.2.0^48^ with automated model selection, ModelFinder^49^ and branch support was estimated using 1,000 replicates of ultrafast bootstrap approximation (UFBoot)^50^.

Lineage nomenclature was defined for ANDV using the M segment as the phylogenetic reference and 1991 as the initial time point, following the Pango lineage nomenclature framework^18^. Lineages were designated for monophyletic groups with high support (UFBoot >=98 and SH-aLRT>=100), and geographic or epidemiological relevance. Putative reassortant lineages were identified based on incongruent phylogenetic topologies across segments and were excluded from the nomenclature as they fall outside the scope of this work (Extended Data Fig.3a-c).

### Bayesian phylogeographic reconstruction

Given the phylogenetic clustering of the MV Hondius outbreak genomes, subsequent time-resolved analyses were restricted to the ANDV lineage (Extended Data Fig.2). The temporal signal was assessed in a regression analysis of the root-to-tip divergence against the tip sampling date in TempEst (v1.5.3)^51^, using the maximum-likelihood tree inferred. Root-to-tip regression plots following dataset refinement are shown in Extended Data Fig.6.

We performed a continuous phylogeographic analysis using the relaxed random walk diffusion model implemented in BEAST X v10.5.0^52^ software package with the BEAGLE^53^ library to improve computation performance. We used a general-time-reversible (GTR) nucleotide substitution model with gamma-distributed among-site rate-heterogeneity. Evolutionary rates were estimated under an uncorrelated relaxed lognormal (UCLD) molecular clock using a constant coalescent tree prior. For each sequence, latitude and longitude were attributed to the most precise district or provincial information available. For the outbreak-associated sequence, geographic location was sampled from a predefined set of candidate locations from the travel history of the index case using a bit-move operator. We ran two independent chains of 100 million generations (sampled every 10,000 states) that were run and then combined to ensure convergence, discarding the initial 10% as burn-in. The chains were then combined with LogCombiner. We assessed convergence using Tracer and reconstructed a maximum clade credibility tree in TreeAnnotator^54^. Effective sample size for all parameters was > 200. The Spread.gl^55^ python package was used to reconstruct the spatiotemporal diffusion patterns.

## Data availability

Sequences are available in the Pathoplexus SeqSet (10.62599/PP_SS_1849.1). Newly generated sequences that have not been previously published will be deposited in Pathoplexus prior to publication. All analysis code and data are available at https://github.com/Liss-UZ/Mv_Hondius_outbreak_project

## Notes

### Competing Interest Statement

The authors have declared no competing interest.

### Author Declarations

Ethics Committee of the Faculty of Medicine of Clinica Alemana Universidad del Desarrollo gave ethical approval for this work.

## REFERENCES

1. WHO Rapid Risk Assessment - Hantavirus outbreak caused by Andes virus, Global v.2. https://www.who.int/publications/m/item/who-rapid-risk-assessment---hantavirus-outbreak-caused-by-andes-virus--global-v.2.

2. Mora-Rillo, M. et al. Andes hantavirus outbreak on a cruise ship—an European Society of Clinical Microbiology and Infectious Disease Emerging Infections Subcommittee rapid assessment. Clin. Microbiol. Infect. 32, 1197–1201 (2026).

3. Andes Hantavirus Outbreak on a Cruise Ship, 2026. N. Engl. J. Med. https://doi.org/10.1056/NEJMc2606496 (2026) doi:10.1056/NEJMc2606496.

4. Complete sequence of Orthohantavirus andesense virus: Swiss resident 2026 - Hantavirus. *Virological* https://virological.org/t/complete-sequence-of-orthohantavirus-andesense-virus-swiss-resident-2026/1023/9 (2026).

5. Preliminary analysis of Orthohantavirus andesense virus sequences from a cruise-ship related cluster, May 2026 - Hantavirus. *Virological* https://virological.org/t/preliminary-analysis-of-orthohantavirus-andesense-virus-sequences-from-a-cruise-ship-related-cluster-may-2026/1029 (2026).

6. Hantavirus outbreak linked to cruise ship travel, Multi-locations. https://www.who.int/emergencies/disease-outbreak-news/item/2026-DON611.

7. Alonso, D. O., et al. Person-to-Person Transmission of Andes Virus in Hantavirus Pulmonary Syndrome, Argentina, 2014. (2020).

8. Martínez, V. P. et al. “Super-Spreaders” and Person-to-Person Transmission of Andes Virus in Argentina. N. Engl. J. Med. 383, 2230–2241 (2020).

9. Martinez-Valdebenito, C. et al. Person-to-Person Household and Nosocomial Transmission of Andes Hantavirus, Southern Chile, 2011. Emerg. Infect. Dis. 20, 1629– 1636 (2014).

10. Sullivan, B. L. et al. eBird: A citizen-based bird observation network in the biological sciences. Biol. Conserv. 142, 2282–2292 (2009).

11. Argentina asiste a países europeos con insumos de diagnóstico para hantavirus. *Argentina.gob.ar* https://www.argentina.gob.ar/noticias/argentina-asiste-paises-europeos-con-insumos-de-diagnostico-para-hantavirus (2026).

12. Palma, R. E., Boric-Bargetto, D., Torres-Pérez, F., Hernández, C. E. & Yates, T. L. Glaciation Effects on the Phylogeographic Structure of Oligoryzomys longicaudatus (Rodentia: Sigmodontinae) in the Southern Andes. PLOS ONE 7, e32206 (2012).

13. Astorga, F. et al. Distributional ecology of Andes hantavirus: a macroecological approach. Int. J. Health Geogr. 17, 22 (2018).

14. Alonso, D. O. et al. Orthohantavirus diversity in Central-East Argentina: Insights from complete genomic sequencing on phylogenetics, Geographic patterns and transmission scenarios. PLoS Negl. Trop. Dis. 18, e0012465 (2024).

15. Shedroff, E. et al. Completed genome segments of Maciel, Lechiguanas, and Laguna Negra orthohantaviruses. Microbiol. Resour. Announc. 13, e00441–24 (2024).

16. Ulloa-Zepeda, L., Parker, E., Pavez, C. & Vial, M. C. Genomic Surveillance of Andes Virus Uncovers Hidden Diversity in Chile. SSRN Scholarly Paper at 10.2139/ssrn.6620250 (2026).

17. Potential geographical and ecological origin of the MV Hondius Andes virus outbreak - Hantavirus. *Virological* https://virological.org/t/potential-geographical-and-ecological-origin-of-the-mv-hondius-andes-virus-outbreak/1039.

18. Colquhoun, R. et al. A generalized lineage nomenclature for viral genomic epidemiology. Nat. Microbiol. 1–10 (2026) doi:10.1038/s41564-026-02468-3.

19. Layan, M. et al. Impact and mitigation of sampling bias to determine viral spread: Evaluating discrete phylogeography through CTMC modeling and structured coalescent model approximations. Virus Evol. 9, vead010 (2023).

20. Hall, M. D., Woolhouse, M. E. J. & Rambaut, A. The effects of sampling strategy on the quality of reconstruction of viral population dynamics using Bayesian skyline family coalescent methods: A simulation study. Virus Evol. 2, vew003 (2016).

21. Gressani, O., Torneri, A., Hens, N. & Faes, C. Flexible Bayesian estimation of incubation times. Am. J. Epidemiol. 194, 490–501 (2025).

22. Charniga, K. et al. Best practices for estimating and reporting epidemiological delay distributions of infectious diseases. PLOS Comput. Biol. 20, e1012520 (2024).

23. Delignette-Muller, M. L. & Dutang, C. **fitdistrplus** : An *R* Package for Fitting Distributions. J. Stat. Softw. 64, (2015).

24. Vial, P. et al. Incubation Period of Hantavirus Cardiopulmonary Syndrome - Volume 12, Number 8—August 2006 - Emerging Infectious Diseases journal - CDC. https://doi.org/10.3201/eid1208.051127 doi:10.3201/eid1208.051127.

25. ANLIS Malbrán identificó hantavirus en roedores de Tierra del Fuego sin relación con el brote del crucero MV Hondius. Argentina.gob.ar https://www.argentina.gob.ar/noticias/anlis-malbran-identifico-hantavirus-en-roedores-de-tierra-del-fuego-sin-relacion-con-el-0 (2026).

26. Minsal. INFORME EPIDEMIOLÓGICO ANUAL, Hantavirus, 2024.

27. Medina, R. A. et al. Ecology, genetic diversity, and phylogeographic structure of andes virus in humans and rodents in Chile. J. Virol. 83, 2446–2459 (2009).

28. Clinical Sequencing Uncovers Origins and Evolution of Lassa Virus: Cell. https://www.cell.com/fulltext/S0092-8674(15)00897-1.

29. Klitting, R. et al. Predicting the evolution of the Lassa virus endemic area and population at risk over the next decades. Nat. Commun. 13, 5596 (2022).

30. Parker, E. et al. Genomics reveals zoonotic and sustained human mpox spread in West Africa. Nature 643, 1343–1351 (2025).

31. Instituto de Salud Pública de Chile. https://www.ispch.gob.cl/biomedico/vigilancia-de-laboratorio/ambitos-de-vigilancia/vigilancia-hanta/.

32. Lessler, J. et al. Incubation periods of acute respiratory viral infections: a systematic review. Lancet Infect. Dis. 9, 291–300 (2009).

33. Czumbel, I. et al. Management and control of communicable diseases in schools and other child care settings: systematic review on the incubation period and period of infectiousness. BMC Infect. Dis. 18, 199 (2018).

34. Czubaj, F. Hantavirus en alta mar: nueva hipótesis sobre el inicio del brote y la pista para llegar a un motorhome clave. LA NACION https://www.lanacion.com.ar/sociedad/hantavirus-en-alta-mar-nueva-hipotesis-sobre-el-inicio-del-brote-y-la-pista-para-llegar-a-un-nid22062026/ (2026).

35. Hjelle, B. et al. Epidemiologic linkage of rodent and human hantavirus genomic sequences in case investigations of hantavirus pulmonary syndrome. J. Infect. Dis. 173, 781–786 (1996).

36. Vial, C. et al. Molecular method for the detection of Andes hantavirus infection: validation for clinical diagnostics. Diagn. Microbiol. Infect. Dis. 84, 36–39 (2016).

37. Chen, S. fastp 1.0: An ultra-fast all-round tool for FASTQ data quality control and preprocessing. iMeta vol. 4 (2025).

38. Nurk, S., Meleshko, D., Korobeynikov, A. & Pevzner, P. A. metaSPAdes: a new versatile metagenomic assembler. Genome Res. 27, 824–834 (2017).

39. Li, H. & Durbin, R. Fast and accurate short read alignment with Burrows-Wheeler transform. Bioinformatics 25, 1754–1760 (2009).

40. Grubaugh, N. D. et al. An amplicon-based sequencing framework for accurately measuring intrahost virus diversity using PrimalSeq and iVar. Genome Biol. 20, 8 (2019).

41. Danecek, P. et al. Twelve years of SAMtools and BCFtools. GigaScience 10, giab008 (2021).

42. Wick, R. rrwick/Porechop. (2026).

43. De Coster, W., D’Hert, S., Schultz, D. T., Cruts, M. & Van Broeckhoven, C. NanoPack: visualizing and processing long-read sequencing data. Bioinformatics 34, 2666–2669 (2018).

44. Li, H. Minimap2: pairwise alignment for nucleotide sequences. Bioinformatics 34, 3094–3100 (2018).

45. Preliminary analysis of Orthohantavirus andesense virus sequences from a cruise-ship related cluster, May 2026 - Hantavirus. *Virological* https://virological.org/t/preliminary-analysis-of-orthohantavirus-andesense-virus-sequences-from-a-cruise-ship-related-cluster-may-2026/1029 (2026).

46. Vecchia, E. D. Pathoplexus: towards fair and transparent sequence sharing. Lancet Microbe 5, (2024).

47. Katoh, K. & Standley, D. M. MAFFT: Iterative Refinement and Additional Methods. in *In*Russell, D. (eds) Multiple Sequence Alignment Methods (ed. Russell, D. J.) vol. 1079 131–146 (Humana Press, Totowa, NJ, 2014).

48. Minh, B. Q. et al. IQ-TREE 2: New Models and Efficient Methods for Phylogenetic Inference in the Genomic Era. Mol. Biol. Evol. 37, 1530–1534 (2020).

49. Kalyaanamoorthy, S., Minh, B. Q., Wong, T. K., von Haeseler, A. & Jermiin, L. S. ModelFinder: Fast Model Selection for Accurate Phylogenetic Estimates. Nat. Methods 14, 587–589 (2017).

50. Hoang, D. T., Chernomor, O., von Haeseler, A., Minh, B. Q. & Vinh, L. S. UFBoot2: Improving the Ultrafast Bootstrap Approximation. Mol. Biol. Evol. 35, 518–522 (2018).

51. Rambaut, A., Lam, T. T., Max Carvalho, L. & Pybus, O. G. Exploring the temporal structure of heterochronous sequences using TempEst (formerly Path-O-Gen). Virus Evol. 2, vew007 (2016).

52. Baele, G. et al. BEAST X for Bayesian phylogenetic, phylogeographic and phylodynamic inference. Nat. Methods 22, 1653–1656 (2025).

53. Ayres, D. L. et al. BEAGLE 3: Improved Performance, Scaling, and Usability for a High-Performance Computing Library for Statistical Phylogenetics. Syst. Biol. 68, 1052–1061 (2019).

54. Rambaut, A., Drummond, A. J., Xie, D., Baele, G. & Suchard, M. A. Posterior Summarization in Bayesian Phylogenetics Using Tracer 1.7. Syst. Biol. 67, 901–904 (2018).

55. Li, Y. et al. spread.gl: visualizing pathogen dispersal in a high-performance browser application. Bioinformatics 40, btae721 (2024).

